# Co-expressed MicroRNAs Associated with An Elevated Psychometabolic Risk Phenotype in Women during Midlife

**DOI:** 10.64898/2026.04.27.26351846

**Authors:** Kayla D. Longoria, Benjamin M. Stroebel, Meghana Gadgil, Nicole Perez, Kimberly A. Lewis, Sandra Weiss, Elena Flowers

**Affiliations:** Department of Physiological Nursing, University of California, San Francisco, CA; Department of Medicine, University of California, San Francisco, CA; Rory Meyers College of Nursing, New York University, New York, NY; UCLA Health, University of California Los Angeles, Los Angeles, CA; Department of Community Health Systems, University of California, San Francisco, CA; Institute for Human Genetics, University of California, San Francisco, CA

## Abstract

**Introduction:** The bidirectional relationship between depression and type 2 diabetes (T2D) is well-established. Women are disproportionately affected by their co-occurrence, particularly during midlife, yet sex- and age-specific studies on phenotypic and mechanistic factors underlying risk for their co-occurrence are limited. The purpose of this study was to identify combined risk profiles (i.e., depression, T2D) in women during midlife and to determine if microRNAs (miRs) that are associated with high-risk profiles provide mechanistic insights into multimorbidity.

**Materials and Methods:** This study included baseline data from women during midlife (ages 40-64 years) who participated in the Diabetes Prevention Program (DPP) (n = 603). Unsupervised k-means clustering was used to identify multimorbid risk profiles. Clinical characteristics included for risk profiling included Beck Depression Inventory (BDI-I), age, BMI, waist circumference, triglycerides, high HDL, FBG, and HbA1c. Associations between risk profiles and individual miRs and principal components of co-expressed miRs were determined via logistic regression models adjusted for participant race and ethnicity. False discovery rate (q< 0.05) was used to control for multiple comparisons.

**Results:** Two distinct profiles were identified, with the high-risk profile characterized by younger age yet higher adiposity, glycemic biomarkers, and depression symptom burden compared to the low-risk profile. MiR-320a and miR-320c were associated with increased odds of high-risk profile assignment, and a co-expression cluster enriched for miRs belonging to the miR-320 family (PC3) was significantly associated with increased odds of high-risk profile assignment. Across all models, Black race demonstrated at least threefold higher odds of high-risk profile assignment.

**Discussion:** These findings highlight distinct multimorbid risk profiles in women during midlife, emphasizing the potential utility of integrated, multidimensional approaches for risk stratification. Findings also revealed mechanisms that may underly risk for co-occurrence of T2D and depression in women during midlife and potential therapeutic targets for prevention and treatment.

## INTRODUCTION

In the United States (U.S.), chronic health conditions are responsible for 90% of the nation’s $4.9 trillion in annual health care expenditures (1). Multimorbidity, defined as having two or more co-occurring chronic conditions without emphasis on an index condition (2), affects approximately one in four U.S. adults and is occurring at younger ages (3). Co-occurring mental and physical health conditions (mixed multimorbidity) are among the most expensive to manage and disproportionately affect women compared to men (4, 5). These sex differences peak during midlife (ages 40-64) but narrow over time, highlighting sexually dimorphic patterns in windows of vulnerability and suggesting factors beyond normative aging processes may contribute to these differences (4, 6). Despite this, multimorbidity research has largely focused on physical morbidities in older adults (age ≥65 years) within mixed-sex cohorts (7), leaving critical gaps in understanding phenotypic and mechanistic factors contributing to women’s risk for co-occurring mental and physical conditions during midlife.

Depression and type 2 diabetes (T2D) are both recognized as leading causes of disability globally and together constitute one of the most extensively studied mixed multimorbidity dyads (8,9). The bidirectional relationship between these conditions is well-documented, with each condition evidenced to increase the risk of developing the other by approximately 30-60% (10, 11). Consistent with temporal patterns observed for mixed multimorbidity, epidemiological studies demonstrate women during midlife are more likely to develop co-occurring depression and T2D than men (4, 12). Midlife marks a distinct life stage for a majority of women, as the menopausal transition initiates biological changes and related symptoms with significant interindividual variability (13). One of the most comprehensive studies on women’s health during midlife showed that this timeframe is a high-risk period for subclinical shifts in clinical characteristics known to increase risk for T2D (e.g., lipids, central adipose (visceral) tissue distribution, insulin resistance) and mental health disturbances (e.g., depression, anxiety) (13–16). However, few studies have examined these biological changes in women during midlife to understand combined T2D and depression risk profiles and related mechanisms that potentially underly this multimorbidity.

MicroRNAs (miRs) are non-coding RNA molecules that regulate protein production by binding to target messenger RNAs (mRNAs) and regulating translation (18). Circulating miRs found in serum and plasma are increasingly recognized as promising biomarkers for mechanisms that contribute to risk for complex conditions, including extrinsic stressors (e.g., behavioral, psychosocial, environmental) (17, 18). Prior studies have independently established associations between circulating miRs and risk for T2D (19–21) and depression (22). Despite the evidence that there may be unique independent and multimorbid risk for these conditions in women during midlife, none of the prior studies have focused specifically on this population. Circulating miRs may be an approach for both identification of women at highest risk for multimorbidity during midlife and provide insights into the mechanisms that underlie this increased risk.

The purpose of this study was to identify multimorbid risk profiles (i.e., depression, T2D) in women during midlife, to determine if miRs are associated with elevated risk, and to provide insights into mechanisms that underlie this multimorbidity. The potential implications of this study are improved risk stratification and identification of potential therapeutic targets to decrease the multimorbidity of T2D and depression in women during midlife.

## METHODS

### Study Design and Participant Data Collection

This study was a secondary analysis of baseline data from a subset of participants in the Diabetes Prevention Program (DPP) trial (23–25). In brief, the DPP trial recruited 3,234 participants between 1996-1999 from 27 centers across the U.S., with oversampling from racially minoritized groups (23). Participants had a body mass index (BMI) >24kg/m^2^, fasting blood glucose (FBG) 95-125 mg/dL, and a 2-hour post-challenge glucose 140-199 mg/dL. Individuals with serious illness or using medications known to alter glucose tolerance were not eligible. At baseline, trained study personnel collected data on demographic characteristics, medical history, depression (Beck Depression Inventory (BDI-I) (26)), as well as measures of glycemic control, body composition, and lipid profiles. Blood was collected by venipuncture into vacutainers containing the stabilizing agent heparin.

The study described in this manuscript included all participants between the ages of 40-64 years who self-reported their sex as female and had baseline data for the clinical characteristics included for risk profiling (i.e., age, FBG, hemoglobin A1c (HbA1c), BMI, waist circumference, high density lipoprotein cholesterol (HDL), triglycerides, BDI-I scores), as well as miR variables (n=603).

### Molecular Data Collection

The Fireplex Multiplex Circulating MicroRNA Assay (Abcam, MA) was used for direct quantification of 58 miRs from plasma collected at the baseline visit of the DPP trial (20). MiRs were hybridized to complementary oligonucleotides covalently attached to encoded hydrogel microparticles. The bound target was ligated to oligonucleotide adapter sequences that serve as universal PCR priming sites. The miR-adapter hybrid models were then denatured from the particles and reverse transcription polymerase chain reaction (RT-PCR) was performed using a fluorescent forward primer. Once amplified, the fluorescent target was rehybridized to the original capture particles and scanned on an EMD Millipore Guava 6HT flow cytometer (Merck KGaA Darmstadt, Germany).

### Statistical Analysis

All statistical analyses were performed using R (version 4.3.2) (27). Descriptive statistics were used to summarize data. Chi-squared (categorical variables) or analysis of variance (ANOVA) (continuous variables) were used to evaluate cluster differences between participant demographic and clinical characteristics.

#### Risk profiling

Risk profile clustering variables were selected based on known T2D risk factors (i.e., age, BMI, waist circumference, triglycerides, high density lipoprotein cholesterol (HDL), FBG, hemoglobin A1c (HbA1c)) and BDI-I scores (21). Unsupervised k-means clustering was conducted using these variables for a range of cluster counts (k=1-10). The average silhouette width, which ranges from -1 to 1 (1 indicating well-matched clusters), was calculated for each K value using the factoextra package in R to assess cluster stability and similarity within each cluster (28). Average silhouette widths supported a K value of two, (average silhouette width 0.19) as best fitting the data. The k=2 model also provided clear delineation between cluster profiles across clinical characteristics. Two-cluster k-means models were subsequently created using the Cluster package in R (29). Clusters were labeled as high and low psychometabolic risk profiles based on mean values of individual clustering variables.

#### MiR normalization and quantification

Expression of individual miRs was normalized using the set of miR probes (i.e., miR-92a-3p, mir-93-5p, miR-17-5p) identified by the geNorm algorithm for each experiment (30). A lower limit of quantification (LLOQ) equal to two time the minimum detectable difference for each miR was applied to the samples, and sub-LLOQ values were filtered. MiRs detected in ≥90% of samples were retained for comparison, then converted into z-scores to account for batch effects between miR data collection timepoints, resulting in 35 miRs retained for analyses.

#### Molecular associations with risk profile assignment

Associations between expression of individual miRs and psychometabolic risk profile assignment were determined via logistic regression models adjusted for participant race and ethnicity. Correction for multiple testing was performed by applying false discovery rate (q < 0.05) (31). Associations between miR PCs and psychometabolic risk profile assignment were determined via a logistic regression model for all PCs adjusted for race and ethnicity.

Principal component analysis (PCA) was performed to reduce the dimensionality of miR data and create a set of uncorrelated principal component (PC) variables reflecting the co-expression of multiple miRs. Selection of PCs to retain for analysis was determined via a visual scree plot analysis. The miRs most important to each PC were determined by the loading coefficients for each PC, in which the ten miRs with the greatest absolute values of their loading coefficients were selected to characterize each PC.

## RESULTS

### Risk profile characteristics

Two distinct risk profiles were identified (**Supplementary Figure 1**). The low-risk profile was older with higher HDL and lower BMI, waist circumference, FBG, HbA1c, triglycerides, and depression symptom burden (**Table 1**). The high-risk profile was younger with lower HDL and higher BMI, waist circumference, FBG, HbA1c, triglycerides, and depression symptom burden (**Table 1**). Risk profiles differed significantly by race and ethnicity, with the low-risk profile being of predominantly White race (65%) and the high-risk profile being of predominantly Black race (36%) (**Table 1**). With the exception of triglycerides, all clinical characteristics demonstrated statistically significant differences between profiles. The primary characteristics distinguishing risk profiles were depression symptom burden (69% higher in the high-risk profile) and measures of body composition (35% higher BMI and 25% higher waist circumference in the high-risk profile).

**Table 1.** Demographic and Clinical Characteristics by Cluster.

| Characteristic<br>mean ± SD or n (%) | Clusters | | | p-value |
| --- | --- | --- | --- | --- |
|  | Overall<br>(n=603) | Low Risk<br>(n=361) | High Risk<br>(n=242) |  |
| Age (years) | 50 ± 9 | 51 ± 10 | 48 ± 9 | <0.0001 |
| BMI kg/m<sup>2</sup>) | 34.9 ± 7.3 | 30.6 ± 3.9 | 41.3 ± 6.4 | <0.0001 |
| Waist (cm) | 103 ± 15 | 94 ± 9 | 117 ± 12 | <0.0001 |
| FBG (mg/dL) | 106 ± 7 | 104 ± 6 | 110 ± 8 | <0.0001 |
| HbA1c (%) | 5.9 ± 0.5 | 5.8 ± 0.4 | 6.1 ± 0.5 | <0.0001 |
| Triglycerides (mg/dL) | 161 ± 90 | 159 ± 92 | 163 ± 86 | 0.5272 |
| HDL (md/dL) | 49 ± 13 | 52 ± 13 | 44 ± 10 | <0.0001 |
| BDI depression score | 4.9 ± 4.4 | 3.9 ± 3.4 | 6.6 ± 5.2 | <0.0001 |
| Race and Ethnicity |  |  |  | 0.0004 |
| White | 357 (59) | 234 (65) | 123 (51) |  |
| Black | 138 (23) | 52 (14) | 86 (36) |  |
| Hispanic | 88 (15) | 58 (16) | 30 (12) |  |
| Other | 20 (3) | 17 (5) | 3 (1) |  |

SD – standard deviation; BMI – body mass index; FBG – fasting blood glucose; HbA1c – hemoglobin A1c; HDL – high density lipoprotein cholesterol; BDI – Beck Depression Inventory

### Individual miRs Associated with Risk Profile Assignment

Logistic regression models adjusted for race and ethnicity identified six individual miRs significantly associated with high-risk profile assignment (**Table 2**) After correction for multiple comparisons, miR-320a-3p (q = 0.0126) and miR-320c (q = 0.0318) remained statistically significant. We also observed that women of Black race had higher odds (odds ratio (OR) >2.0, p<0.0001 for all) for high-risk profile assignment in models for all six miRs (**Table 2**).

**Table 2.** Individual MiRs Associated with Odds of High Psychometabolic Risk Assignment.

| **Variable** | **OR** | **95% CI** | **p-value** | **FDR** |
| --- | --- | --- | --- | --- |
| **miR-27a-3p** | 0.84 | 0.71, 0.99 | **0.0354** | 0.2063 |
| **Black** | 3.35 | 2.22, 5.11 | **<0.0001** |  |
| **Hispanic** | 1.01 | 0.61, 1.65 | 0.9565 |  |
| **Other** | 0.32 | 0.07, 0.97 | 0.0724 |  |
| **miR-320c** | 1.29 | 1.10, 1.53 | **0.0018** | **0.0318** |
| **Black** | 3.33 | 2.21, 5.05 | **<0.0001** |  |
| **Hispanic** | 1.05 | 0.64, 1.72 | 0.8374 |  |
| **Other** | 0.33 | 0.08, 1.03 | 0.0863 |  |
| **miR-93-5p** | 0.83 | 0.69, 0.98 | **0.0296** | 0.2063 |
| **Black** | 3.12 | 2.08, 4.72 | **<0.0001** |  |
| **Hispanic** | 0.98 | 0.60, 1.60 | 0.9503 |  |
| **Other** | 0.35 | 0.08, 1.08 | 0.1038 |  |
| **miR-17-5p** | 0.79 | 0.66, 0.94 | **0.0076** | 0.0891 |
| **Black** | 3.22 | 2.14, 4.88 | **<0.0001** |  |
| **Hispanic** | 0.97 | 0.59, 1.58 | 0.9047 |  |
| **Other** | 0.36 | 0.08, 1.10 | 0.1091 |  |
| **miR-22-3p** | 1.29 | 1.07, 1.59 | **0.0106** | 0.0931 |
| **Black** | 3.00 | 2.0, 4.55 | **<0.0001** |  |
| **Hispanic** | 0.90 | 0.54, 1.48 | 0.6936 |  |
| **Other** | 0.30 | 0.07, 0.93 | 0.0608 |  |
| **miR-320a-3p** | 1.35 | 1.15, 1.60 | **0.0004** | **0.0126** |
| **Black** | 3.29 | 2.18, 5.0 | **<0.0001** |  |
| **Hispanic** | 1.03 | 0.62, 1.69 | 0.8964 |  |
| **Other** | 0.31 | 0.07, 0.96 | 0.0699 |  |

**Legend.** False discovery rate was calculated based on all 35 included miRs. White race was used as the reference group.
OR – odds ratio; CI – confidence interval; FDR – false discovery rate

### Co-Expressed MiRs Associated with Risk Profile Assignment

Scree plot analysis identified four distinct PCs of co-expressed miRs (PC1-PC4), which explained 14% of the total variance and were retained for further analysis (**Supplementary Figure 1**). In the logistic regression model adjusted for race and ethnicity that included all four miR PCs, PC3 was significantly associated with increased odds of high-risk profile assignment (OR 1.22, 95% CI 1.10, 1.36, p = 0.0032). The top 10 miRs loading PC3 included miR-320c, miR-320a-3p, miR-126-3p, miR-486-3p, miR-93-5p, miR-17-5p, let-7c-5p, let-7f-5p, miR-30a-5p, and miR-122-5p. Additional miR loading coefficients for all PCs are shown in **Supplementary Table 1**. Similar to what was observed for individual miRs, in the co-expressed miRs model, women of Black race had over three times greater odds of being assigned to the high-risk profile compared to women of white race (OR 3.36, 95% CI 2.19, 5.20, p < 0.0001), after adjusting for all four PC co-expression clusters.

## DISCUSSION

By integrating data commonly collected in clinical practice for screening for T2D and depression, we identified two distinct psychometabolic risk profiles in women during midlife. Profiles suggest metabolic risk factors and subclinical depression may co-occur in clinically meaningful ways that heighten risk for multimorbidity and potentially accelerate disease progression yet remain largely unrecognized until symptoms have escalated to a point of clinical significance (21). These findings also highlight the potential value of multidimensional risk assessments that may enhance precision in risk stratification for early intervention more targeted prevention and treatment strategies.

The high-risk profile, despite being younger (48 years versus 52 years), exhibited significantly elevated risk across nearly all metabolic risk factors and had a higher depression symptom burden. This pattern may reflect a biobehavioral feedback loop, where physiological shifts that occur during midlife converge with depression symptoms that influence health-related behaviors (e.g., nutrition, physical activity), contributing to a more adverse metabolic risk profile despite being younger in age (13). Additionally, while this study was not designed to parse out the impact of psychosocial and environmental exposures (e.g., systemic oppression), consistently higher odds of high-risk profile assignment among women who were categorized as Black aligns with broader evidence of higher chronic disease burden among historically marginalized populations (32–34), including women during midlife (35). Taken together, these findings suggest potential patterns of clustering or risk factors and highlight how subclinical interactions across T2D and depression risk domains may compound multimorbidity and potentially accelerate disease progression.

Prior studies indicate miRs are biomarkers for incident T2D and glycemic progression (19, 20, 36), as well as longitudinal changes in relation to established T2D risk factors over time (37). The current study is the first to characterize miRs associated with mixed multimorbidity (i.e., T2D, depression) in women during midlife, who may be at increased risk for both of these conditions due changes that occur during this life stage (13–16). We identified both individual and co-expressed miRs associated with increased psychometabolic risk, some of which have existing known mechanistic relationships with risk for T2D (38) and/or depression (39, 40). Among individual miRs identified, miR-320a and miR-320c emerged as the strongest and most consistent miRs associated with high-risk profile assignment. The miR-320 family has been implicated in multiple processes related to risk for T2D, including insulin resistance, diabetic cardiomyopathy, adipocyte function, and inflammation (38). Our own prior studies have shown these miRs are associated with changes in weight over time (37), response to metformin for blood glucose control (41), and target pathways related to metabolism and inflammation (42). These miRs have also been linked to depression (22), including decreased levels of circulating miR-320a associated with new onset depression (43). These findings support a potential role for miR-320a and miR-320c as predictors of risk for co-occurring depression and T2D during midlife in women and as potential therapeutic candidates.

Because miRs exhibit coordinated co-regulation of target mRNAs that impact multiple biological pathways, we also evaluated co-expressed miRs in PC3. MiR-320a and miR-320c most strongly loaded on the co-expression cluster (PC3) associated with high-risk profile assignment. In addition, two of the miRs (miR-17, miR-93) that were individually associated with the high-risk profile (p<0.05) were among the top 10 miRs to load on PC3. Among the top 10 miR loading coefficients for PC3, the remaining three miRs (miR-126, miR-486, miR-7 family) have all previously been reported in studies focused on T2D (36, 37, 44, 45), though none were reported in a systematic review of miRs and depression (22). These observations provide multiple angles of statistical evidence for a subset of miRs that may underly the co-occurrence of depression and T2D in women during midlife.

In models that assessed for individual and combined associations between miRs and psychometabolic risk, Black race was significantly associated with high-risk profile assignment. Prior studies have described sociodemographic differences in body composition, some with linkage to geographic ancestry (19, 20, 36, 42) and others to psychosocial factors including racism (46–48). While we hypothesize that miRs may be an approach to identify multidimensional indicators of risk, this study was focused on identifying potential mechanisms underlying the co-occurrence of depression and T2D. Therefore, these incidental findings warrant further investigation, as broader evidence suggests structural and contextual factors that shape lived experiences may explain biological differences between race and ethnic groups (49).

### Limitations

This was a secondary analysis and therefore limited to the variables and data collected in the DPP trial. Depression symptoms were assessed using the BDI-I version that was available at the time of data collection, which may limit direct comparability with the revised BDI used currently (BDI-II). This study did not have quantitative information about the menopausal transition, preventing us from knowing whether participants were in the pre-, peri-, or post-menopause state. However, the menopause transition occurs over the course of years and will manifest different symptoms at different times within individuals. It is likely that increased risk for T2D and depression occurs on a continuum throughout the age range included in this sample (40-64 years). Historic limitations on definitions of race and ethnicity as well as participant privacy concerns limited reporting in the DPP sample, with race and ethnicity being collapsed into a single, four-level variable. In the absence of an estimate of genetic ancestry, the observed associations between risk profile and Black race cannot differentiate between risk potentially due to genetic ancestry versus social exposures versus both.

## CONCLUSION

This study identified miRs, both singularly and based on co-expression patterns, that are associated with multimorbidity of T2D and depression in women during midlife. While there is strong evidence to suggest bidirectional relationships between T2D and depression, most studies have taken place in older adults and have not evaluate multimorbid risk profiles and potentially shared mechanisms. We identified differences in risk profiles among middle-aged women with prediabetes and provide evidence that miRs may be biomarkers to for identification of women at greatest risk for mixed multimorbidity during midlife. The potential clinical implications include earlier detection of multimorbid risk and particularly during midlife, future identification of therapeutic targets regulated by miRs associated with multimorbidity, and optimization of prevention and treatment interventions. Given that T2D and depression are among the most expensive morbidities to manage, which may be compounded when occurring together, there is also the potential for decreased costs related to disease prevention and management.

## STATEMENTS

### Parent trial registration

NCT00004992

### Data availability statement

Data analyzed for the current study are publicly available through the NIDDK data repository and through the Harvard Dataverse repository: https://doi.org/10.7910/DVN/MJDBJE.

### Ethics statement

The study was approved by University of California, San Francisco and conducted in accordance with the local legislation and institutional requirements. The participants provided their written informed consent to participate in this study.

### Author Contributions

KDL: conceptualization, methodology, formal analysis and writing of original draft. BMS: methodology, conducting the formal analysis, and writing – review & editing. MG, NP, KAL, SW: writing – review & editing. EF: supervision of conceptualization, methodology, and formal analysis; writing – review & editing.

### AI disclosure

No Generative AI was used in the preparation of this manuscript.

