## Supplementary figures and images for "Co-expressed MicroRNAs Associated with An Elevated Psychometabolic Risk Phenotype in Women during Midlife"

### Supp Table 1

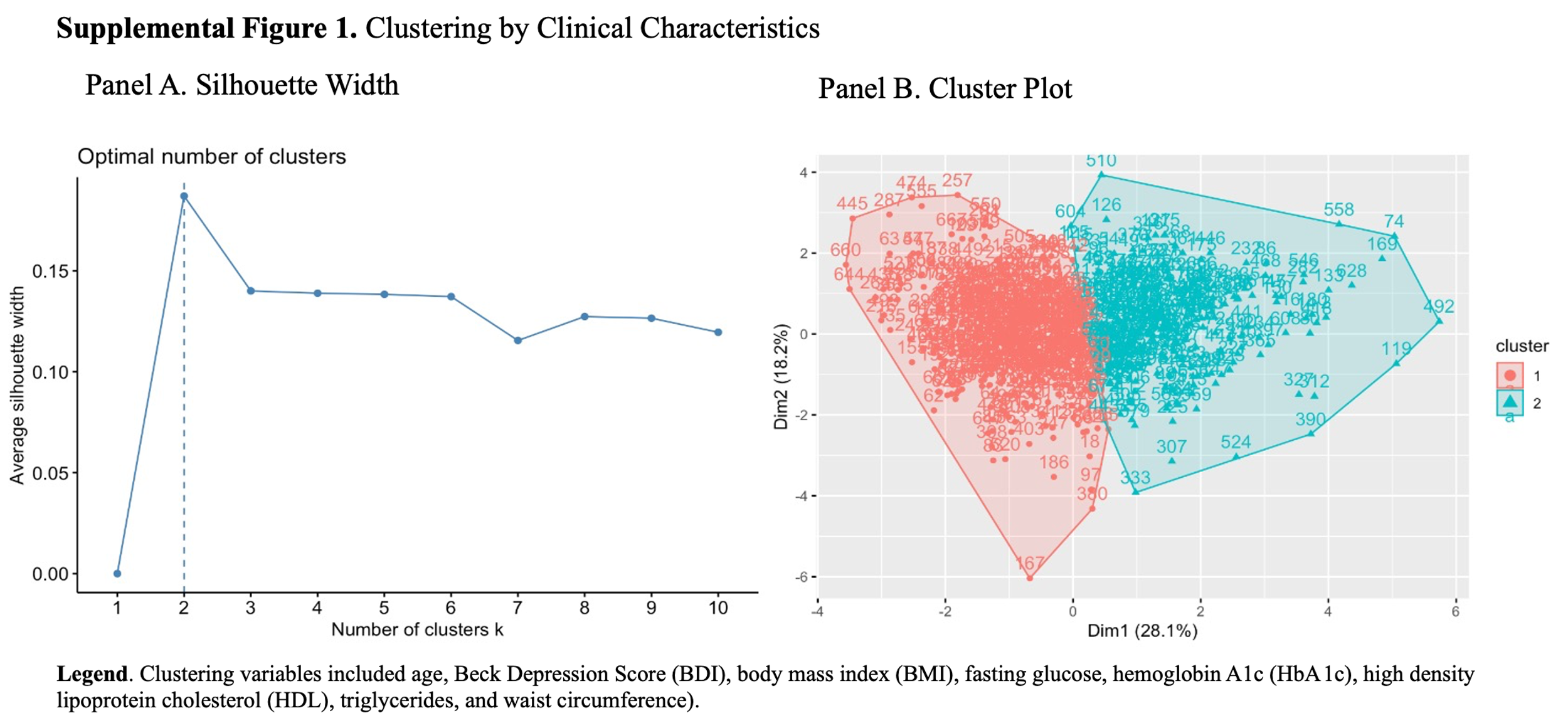
