## Supplementary material for "Co-expressed MicroRNAs Associated with An Elevated Psychometabolic Risk Phenotype in Women during Midlife": Supp Table 2

The *Homo sapiens* microRNA-target interaction (MTI) database was downloaded from miRTarBase. We used R to filter the mRNA gene targets of the identified miRs. Filters for the search included strong, experimentally validated (i.e. Western blot, qRT-PCR, and reporter assay) evidence. Within the OrganismsDbi v1.36.0, we used the Homo.sapiens package (v1.3.1) to determine the entrez IDs for target mRNAs, which were added to results from miRTarBase.
